# QT Interval Prolongation in Patients Treated for COVID-19

**DOI:** 10.1101/2020.12.10.20246975

**Authors:** Parham Habibzadeh, Abdollah Sarami, Mahboobeh Yadollahie, Kourosh Hashemiasl, Arezoo Salahi, Farrokh Habibzadeh

## Abstract

**Background:** Many of the drugs commonly used for the treatment of COVID-19 cause QT interval prolongation and increase the risk of life-threatening cardiac arrhythmias. It has been shown that maintaining serum potassium and magnesium levels above 4 and 3 mg/dL, respectively, would prevent the QTc prolongation.

**Objective:** To determine if keeping only the serum magnesium level above 3 mg/dL could be considered an effective measure to prevent QTc prolongation in patients with COVID-19 receiving these drugs.

**Methods:** In a retrograde observational study, QTc interval was measured in 14 patients diagnosed with COVID-19 before and 3 days after initiation of treatment with either hydroxychloroquine or lopinavir-ritonavir, while their serum magnesium levels were kept ≥3 mg/dL.

**Results:** The baseline QTc interval of 412 (SD 36) ms significantly increased by an average of 34 (95% CI 13 to 55) ms after 3 days of treatment. 5 patients, mostly those with lower serum potassium levels, had QTc prolongation ≥60 ms.

**Conclusion:** Although it seems that the risk of fatal cardiac arrhythmias in this setting is not high, it is prudent to monitor the serum electrolytes, particularly potassium, in patients with COVID-19 who are treated with either hydroxychloroquine or lopinavir-ritonavir.

## Introduction

The novel coronavirus, SARS-CoV-2, infection has now become a major global concern (1). No effective treatments or vaccines exist for COVID-19. Nor do we know much about the recovery of patients. Some researchers report a high re-positive RT-PCR test among patients recovered from the disease (2, 3). The patients can potentially transfer the disease to another person after a median of almost 20 days of complete resolution of their symptoms. Therefore, empirical treatments have widely been used around the globe in order to combat this infection. Many have tried various drugs with a wide range of side effects. Hydroxychloroquine and lopinavir/ritonavir (Kaletra^®^) are among the drugs commonly used. However, these drugs are well-known to cause QT interval prolongation and increase the risk of *torsades de pointes*, a life-threatening ventricular arrhythmia resulting in sudden cardiac death (4, 5). Stress, fever, and electrolyte imbalance are among other factors making these patients prone to arrhythmias.

A recent study has shown that maintaining serum potassium and magnesium levels above 4 mEq/L and 3 mg/dL, respectively, during the treatment of patients with COVID-19 infection, would prevent QTc prolongation in a series of 13 patients treated with one or a combination of hydroxychloroquine, lopinavir/ritonavir, and azithromycin (6). Another study on 13 patients with COVID-19 not being monitored for serum electrolytes, shows that treatment with hydroxychloroquine and/or azithromycine does not significantly prolong the QTc interval (7). We thus conducted this study to determine whether keeping only the serum magnesium level above 3 mg/dL could be considered an effective measure to prevent QTc prolongation in patients with COVID-19 treated with these medications.

### Patients and Methods

In a retrospective observational study, we tried to detect a difference of 30 ms or more in pairs of QTc intervals measured before and 3 days after the initiation of the treatment in a group of patients with COVID-19. Considering an estimated SD of 22 ms in QTc, an acceptable type I error of 0.05, a study power of 0.95, and an estimated correlation coefficient of 0.3, the estimates came from a previous study (6), we came to a minimum sample size of 10 patients (pairs of data).

We studied 14 (9 male and 5 female) patients randomly selected from our patients with mild to moderate COVID-19 admitted in May 2020 to the Great Oil Hospital, Ahwaz, now an epicenter of the disease in southwestern Iran. The diagnosis of COVID-19 was made by RT-PCR on nasopharyngeal samples taken from the patients, according to a method described earlier (8). The patients were treated with either hydroxychloroquine (a loading dose of 400 mg po q12h for one day, followed by 200 mg po q12h) or lopinavir/ritonavir (400/100 mg po q12h). They received magnesium supplementation to keep their serum magnesium above 3 mg/dL.

### Measurement of the QTc Interval

A 12-lead ECG was taken from each patient on admission and 3 days after the initiation of the treatment. The QT interval was measured from the onset of the Q wave, or R wave if no Q wave was identified, to the end of the T wave. The end of the T wave was found using the “tangent” method (9)—the pint was defined as the intersection between the isoelectric line and a line tangent to the steepest last limb of the presumed T wave. QTc was then calculated from the QT and RR intervals using the Bazett’s formula (10). QTc prolongation was defined as QTc ≥500 ms or QTc prolongation ≥60 ms compared to the baseline value (11).

### Ethics

The study protocol was approved by the Petroleum Industry Health Organization Institutional Review Board. Informed written consents were taken from all study participants. This study was conducted in accordance with the ethical principles outlined in the Declaration of Helsinki. The authors are accountable for all aspects of the work in ensuring that questions related to the accuracy or integrity of any part of the work are appropriately investigated and resolved.

### Statistical Analysis

*R* software ver 4.0.2 (2020-06-22) and IBM^®^ SPSS^®^ Statistics for Windows^®^ ver 20 were used for data analysis. One-sample Kolmogorov-Smirnov test was used to check if a continuous variable had a normal distribution. Normally distributed continuous variables were presented as mean (SD). *Student’s t* test for paired data was used to measure the mean difference between QTc intervals measured before and after initiation of the treatment. *R* software was used for drawing the box and whisker plot.

## Results

The 14 studied patients had a mean age of 63 (SD 13, range 36 to 84) years. Nine patients had comorbid conditions on admission—6 had diabetes mellitus; 4, ischemic heart disease; and 3, hypertension. They had a mean serum potassium and magnesium levels of 4.3 (SD 0.6) mEq/L, and 2.0 (SD 0.3) mg/dL, respectively, on admission.

The patients had a mean baseline QTc interval of 412 (SD 36) ms, prior to start of any treatments. The QTc measured 3 days after initiation of the treatment, significantly increased by an average of 34 (95% CI 13 to 55) ms (Fig. 1). None of the patients developed a QTc interval ≥500 ms. However, the QTc prolongation was ≥60 ms in 5 patients (orange line segments, Fig. 1). The mean baseline serum potassium level for these patients (3.9, SD 0.5 mEq/L) was 0.66 (95% CI 0.01 to 1.32) mEq/L, on average, lower than that in those who did not develop a clinically significant QTc prolongation. There was no incidence of arrhythmias or sudden cardiac death among the patients studied.

**Figure 1:**
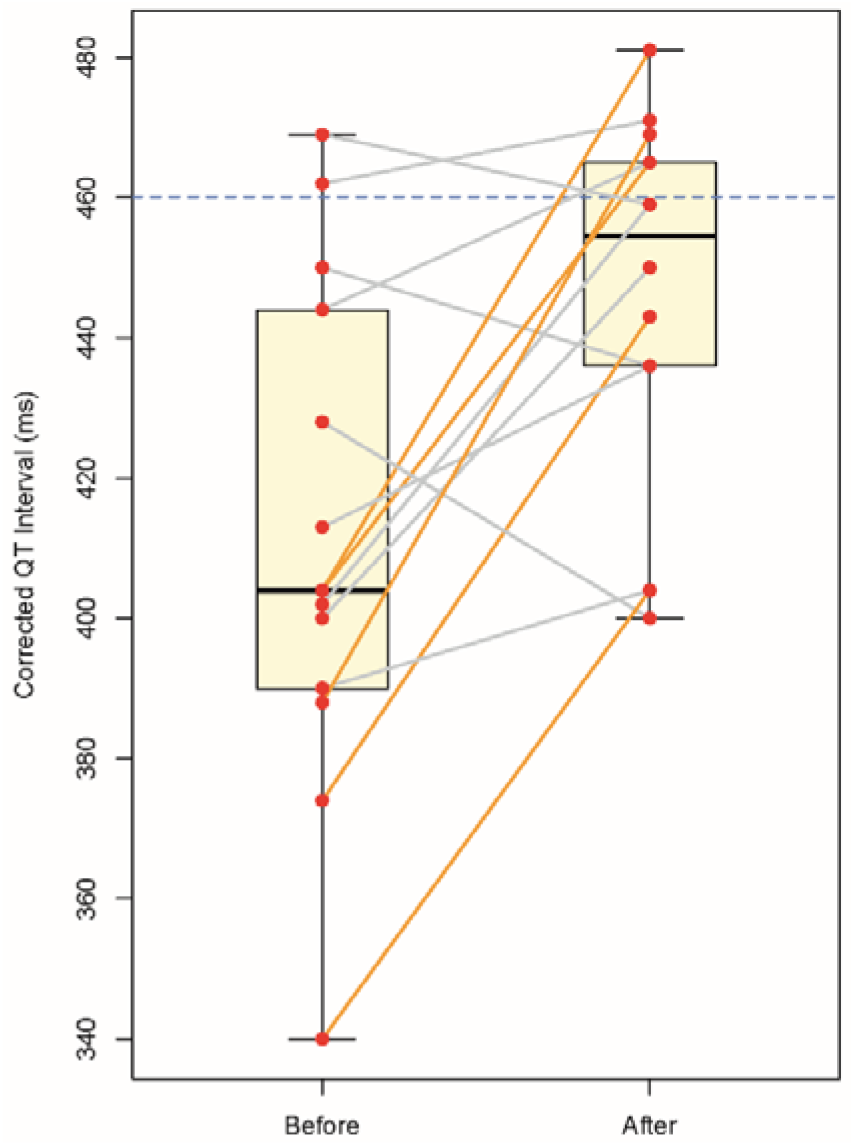
Box and whisker plot of corrected QT interval before and after treatment in patients diagnosed with COVID-19 infection. Orange line segments represent an increase in QTc interval ≥60 ms. Horizontal blue dashed line represents the upper normal limit of the QTc interval (460 ms) (13).

## Discussion

We found that by only keeping their level of serum magnesium levels above 3 mg/dL, one-third of patients with COVID-19—mostly those with a lower normal serum potassium— developed clinically significant QTc prolongation after the initiation of treatment with either hydroxychloroquine or lopinavir/ritonavir. Another study on 13 patients diagnosed with COVID-19 who had a mean baseline QTc interval similar to that of our patients, shows that close monitoring of both serum potassium and magnesium levels would prevent a significant change in QTc intervals as a result of the medications used in the treatment of COVID-19 with known QTc interval prolonging properties (6). This finding would underline the importance of monitoring and correcting serum potassium levels in addition to serum magnesium. Hydroxychloroquine is known to cause QTc prolongation by blockade of ether-à-go-go-related gene (HERG) potassium channel, which can result in fatal ventricular arrhythmias (12, 13). Lopinavir and ritonavir can also cause dose-dependent blockade of HERG potassium channels (14). This might explain our findings in this study and highlights the importance of close monitoring of serum potassium in this setting.

One of the limitations of our study was its observational nature and low sample size. However, despite the low sample size, our design had more than 95% power to detect an increase in QTc interval of at least 60 ms.

A clinically significant QTc prolongation was found in one-third of studied patients. Based on our findings, although it seems that the risk of fatal cardiac arrhythmias in this setting is not high, it is prudent to monitor the serum electrolytes, particularly potassium, in light of the observed QTc prolongation.

## Data Availability

Raw data are available from the corresponding author, if reasonable reasons are provided.

## Data Availability

Raw data are available from the corresponding author, if reasonable reasons are provided.

